# Inflammatory cytokines and a diverse cervicovaginal microbiome are associated with cervical disease in Hispanics living in Puerto Rico

**DOI:** 10.1101/2023.04.06.23288256

**Authors:** Eduardo Tosado-Rodríguez, Loyda B. Mendez, Ana M. Espino, Stephanie Dorta-Estremera, Edna E. Aquino, Josefina Romaguera, Filipa Godoy-Vitorino

## Abstract

Cervical cancer (CC) is women’s fourth most common cancer worldwide. A worrying increase in CC rates in Hispanics suggests that besides Human Papillomaviruses infections, there may be other biological causes promoting the disease. We hypothesized that the cervical microbiome and the epithelial microenvironment favoring inflammation is conducive to disease progression. There is a lack of studies examining the part played by microbial populations in the development of precancerous lesions into cancer in Hispanic women. We aimed to investigate the relationship between the cervicovaginal microbiome and inflammation in Hispanic women living in PR while considering cervical neoplasia and HPV infection. Cervical samples collected from 91 participants coming to clinics in San Juan, underwent 16S rRNA genes (V4 region) profiling, and cytokines were measured using Luminex MAGPIX technology. Cytokines were grouped as inflammatory (IL-1β, TNFα, IFNγ, IL-6), anti-inflammatory (IL-4, IL-10, TGFβ1), and traffic-associated (IL-8, MIP1a, MCP1, IP10). They were related to microbes via an inflammation scoring index based on the quartile and tercile distribution of the cytokine’s concentration. We found significant differences in the diversity and composition of the microbiota according to HPV risk, cervical disease, and cytokine abundance. The most dominant community state type (CST) was CST IV with ∼ 90% dominance in participants with high-grade squamous intraepithelial lesions and high-risk HPV. The increasing concentration of pro-inflammatory cytokines was associated with a decrease in *L. crispatus*. In contrast, dysbiosis-associated bacteria such as *Gardnerella*, *Prevotella*, *Atopobium* concomitantly increased with pro-inflammatory cytokines. Our study highlights that the cervical microbiota of Hispanics living in PR is characteristically diverse, regardless of HPV status and that dysbiosis associated with a decrease in *Lactobacillus* marks inflammatory processes. Therefore, the joint host-microbe interaction analyses via cytokine signaling and microbiota in precancerous lesions is confirmed to have great translational potential.

## Introduction

Cervical cancer (CC) is the 4th most common cancer among women and the 4th leading cause of cancer death in females worldwide (Cohen et al. 2019). Although CC rates have decreased in the USA (Chatterjee et al. 2016), the incidence and mortality from cervical cancer mainly occurs in low resource settings (∼ >85%) (Ferlay et al. 2010). According to the Pan American Health Organization for 2012, 80% of American deaths due to CC occur in Latin America and the Caribbean (1). Puerto Rico (PR) has the highest age-adjusted incidence of CC in the US (Ortiz et al. 2010), with notably higher incidence from 2001 to 2017, increasing from 9.2 to 13 per 100,000 person-years (Ortiz et al. 2021). PR also has a 79.3% CC screening rate that does not achieve CDC’s screening recommendations of 93% (Centers for Disease and Prevention 2012; Ortiz et al. 2010) posing a strain on health systems in the island (2).

Persistent infection with oncogenic Human Papillomavirus (HPV) is the most important cause of CC. HPV infections range from 90–100% prevalence in cervical abnormalities, from high-grade squamous intraepithelial lesion -HGSIL-to severe dysplasia and cancer (3), where the viral genome integration modulates the host cell cycle promoting neoplasia progression (4,5). However, other factors influence cervical disease progression, including lifestyle, genetic predisposition, immunosuppression, early onset of sexual activities, and many pregnancies and deliveries (6–8). Nevertheless, why only a fraction of women acquires a long-lasting, persistent high-risk HPV (HR-HPV) that puts them at risk for developing Cervical Intraepithelial Neoplasia (CIN) remains unknown. One potential explanation for this variation could be the cervicovaginal microbiota and its related processes. Recent studies suggest an association between certain bacterial community types of the vaginal microbiota, HPV infection, and cervical disease (9–11). Additionally, processes like bacterial vaginosis, cervical inflammation, and increased vaginal pH are all known to affect susceptibility to cervical HPV (12).

Women of different ethnic groups differ in their vaginal microbiome signatures (13). The cervicovaginal microbiome has been characterized into five different community state types (CST) according to the dominance of *Lactobacillus* species, a concept that was introduced in 2011 by Ravel et al. to categorize vaginal microbial communities among reproductive-age women (13). CST-I refers to communities dominated by *L. crispatus* (CST-I), CST-II by *L. gasseri* , CST-III by *L. iners*, CST-V by *L. jensenii* (CST-V), or non-*Lactobacillus* dominated and highly diverse profiles such as CST-IV. Furthermore, CST-IV is often characterized by three sub-CSTs: 1) CST IV-A has a high relative abundance of Candidatus *Lachnocurva vaginae* and a moderate relative abundance of *G. vaginalis* and *Atopobium vaginae*, 2) CST IV-B also has a moderate relative abundance of *G. vaginalis* and *Atopobium vaginae,* while 3) CST IV-C has low relative abundance of *Lactobacillus* spp., as well as low levels of *G. vaginalis, A. vaginae*, and *Ca. L. vaginae* with high facultative and strictly anaerobic bacteria (14). While *Lactobacillus* dominates the cervicovaginal microbiome of Caucasians, Hispanic and black women have higher diversity and reduced *Lactobacillus* populations (13). A previous study demonstrated that Hispanics living in PR have a different microbial profile than Caucasians, and that patients with high-risk HPV and CIN3 cervical lesions had dominant *Atopobium vaginae* and *Gardnerella vaginalis* (11,15). However, these results could have been improved, as they identified microbiota using cervical swabs, failing to address the host response. Recent studies have highlighted the benefit of collecting cervical lavages to understand the extent of the dynamic host-microbe symbioses via proteomic, metabolomic, or cytokine profiling analyses (16–19).

We hypothesized that there would be changes in the cervical microenvironment with cervical disease and HPV risk and that markers of inflammation would be associated with a decrease in *Lactobacillus* and overall dysbiosis. To test this hypothesis, we investigated the relationship between the cervicovaginal microbiome with cervical neoplasia and HPV infections and characterized the host immune profiles through profiling cytokine expression analyses from cervical lavages.

This study provides insight into specific bacterial taxonomic changes promoting a pro-inflammatory state associated with cervical dysplasia and represents the first effort to combine microbiome and cytokine profiling in Hispanics living in Puerto Rico.

## Materials and Methods

### Participant Recruitment and Sample Collection

Women coming for routine medical care to the University of Puerto Rico and San Juan City clinics (San Juan Metropolitan area), were recruited as part of IRB protocol 1050114 with IBC protocol 94620. Only women who did not meet the exclusion criteria were recruited and evaluated. Exclusion criteria included: 1) Active urinary infections, 2) history of common urinary incontinence, 3) sexually transmitted diseases (STDs), 4) antibiotics taken in the prior month, 5) candidiasis, and 6) treatment for or suspicion of prior toxic shock syndrome. All subjects were informed of the study’s sampling procedure, risks, and benefits with written informed consent and a verbal explanation of the document. A detailed questionnaire from which some of the metadata variables were collected was administered. All procedures were done according to the Declaration of Helsinki. The date range in which human subjects’ data/samples were collected was between November 2017 and February 2020. The study and its procedures were conducted from March 2020 and October 2022.

A total of 91 women, 21-60 years old and able to provide informed consent, were enrolled in our study. Cervical samples were collected through speculum access during the gynecological examination by using cotton tip collection swabs. Sterile Catch-All Specimen Collection Swabs (Epicentre Biotechnologies, Madison WI, United States) were placed in the cervix, rotated along the lumen with a circular motion, and then placed in sterile 2 milliliters (mL) microtubes with cap. We collected approximately 10mL of cervical lavages (CVL) by injecting pure certified nuclease-free water (Growcells, Irvine, CA) into the vaginal canal. Collected lavages were stored in a clean 15mL collection tube and pH was measured with hydrion wide range pH paper strips (Merck, Germany). All samples were stored at −80◦C and processed for nucleic acid extraction and cytokine profiling at our UPR School of Medicine laboratory to reduce processing variation.

### Cytokine Detection and Measurement with Bio-Plex Immunoassay System

A total of 91 cervical lavages were tested in duplicate with a multiplex solid-based immunoassay (Bio-Plex Pro Human Cytokine 10-plex Assay, Bio-Rad) using the same volume of CVL across samples (80 uL) for the Luminex MAGPIX® system. Cytokines/chemokines analyzed were IL-1β, IL-4, IL-6, IL-8, IL-10, TNF-α, IFNγ, IP10, MIP1, and MCP1. Also, 91 samples were tested in duplicate using Bio-Plex Pro TGF-β 3-plex immunoassay (BioRad, Hercules, CA, USA). We performed all analyses following the manufacturer’s recommendations.

### Cytokine/Chemokine Profile Analyses

Data were analyzed with the Bio-Plex Manager 6.1 software (BIO-RAD, Hercules, CA) using a 5-parameter logistic curve that was verified for normality before statistical testing (20). Inflammation scores were assigned to each cytokine/chemokine for each sample using Fluorescent Intensity (FI) results. Scores were calculated by distributing samples across statistical quartiles or tertiles. If a sample was within the first quartile, a score of 1 was assigned, that is for all other three quartiles (score 1 < 0.25 < score 2 < 0.50 < score 3 < 0.75 < score 4 < 1). TGF-β scores were calculated using tertiles with scoring categories including: score 1 < 0.33, score 2 from 0.33 to 0.66 and score 3 < 1. Cytokines were grouped according to their function on host physiology, including: pro-inflammatory (IL-1β, TNF-α, IFNγ, IL-6), anti-inflammatory (IL-4, IL-10, TGFβ1) and traffic-associated cytokines(IL-8, MIP1a, MCP1, IP10) (21,22). With the scoring in either tertiles or quartiles, we also created a categorical variable, to which scores of 2 or 1 corresponded to low levels of cytokines and > 2 high levels. These ranked score assignments using the distribution of cytokines were directly related to the concentration (fluorescence intensity) of cytokines/chemokines in a sample. Therefore, we used them to combine microbial community analysis (16S rDNA profiles) with inflammation scores as a metadata variable (see methods section “Statistical Analyses of the Microbiota Profiles” for more details). This combination allowed us to detect specific taxa associated with altered cytokine profiles. For comparisons between multiple groups, we used ANOVA, and for comparisons between two groups, we used the student’s *t*-test. All statistical analyses were done using GraphPad Prism 6.0 (San Diego, CA).

### DNA Extraction and 16S rDNA Sequencing

Genomic DNA extractions from swabs were performed with the Qiagen Power Soil Kit (QIAGEN LLC, Germantown Road, Maryland, USA) following an optimized protocol for these types of samples as described in a previous study (11). DNA was quantified with a Qubit® dsDNA HS (High Sensitivity) Assay (Waltham, Massachusetts, US) (ranging from 5-100ng/µL). An average of 10-30ng/µL genomic DNA was shipped to an outsourced sequencing facility.

DNA from cervical samples was normalized to 4nM during 16S library prep. The V4 hypervariable region of the 16S ribosomal RNA marker gene (∼291bp) was amplified using universal bacterial primers: 515F (5’GTGCCAGCMGCCGCGGTAA3’) and 806R (5’GGACTACHVGGGTWTCTAAT3’) as in the Earth Microbiome Project (http://www.earthmicrobiome.org/emp-standard-protocols/16s/) (23). The 16S-rRNA reads and corresponding sample metadata was deposited in QIITA study ID 12871 (24), and the raw sequences are available in the European Nucleotide Archive ENA Project under accession number EBI: ERP136546.

### HPV Genotyping process

We used a kit based on a highly sensitive short-polymerase chain reaction-fragment assay (Labo Biomedical Products, Rijswijk, The Netherlands, licensed Innogenetics technology). It uses SPF10 primers to amplify a 65-bp fragment of the L1 open reading frame of HPV genotypes, followed by a Reverse-Hybridization step to determine which HPV type is present by comparing to kit-provided controls. This assay allows for the identification of the following common mucosal HPV genotypes: 14 High-risk types including (HPV 16, 18, 31, 33, 35, 39,45, 51, 52, 56, 58, 59, 66 and 68/73) and 11 Low-risk types (6, 11, 34, 40,42, 43, 44, 53, 54,70, 74). Previous studies explain the method in detail (Vargas-Robles et al. 2018)Godoy-Vitorino et al. 2018).

### Quality Control and Sequence Processing

Raw read pre-processing of demultiplexed files was done with a Phred offset of 33 and default parameters using split libraries FASTQ (QIIMEq2 1.9.1) in QIITA (24). Sequences were trimmed to 250 bp, followed by the deblurring workflow (deblur 1.1.0) (24,25). For taxonomy assignment we used a modified Greengenes reference database (Greengenes_13.8) (26,27) with a minimum similarity threshold of 97%. The species table was downloaded for downstream analyses using a locally run version of QIIME (28). Singletons, amplicon sequence variants (ASVs) with less than three reads, and sequences matching chloroplasts, mitochondria, and taxonomically unassigned sequences were removed from downstream analyses. All samples were rarefied to 7400 reads per sample.

### Vaginal Community State Type Nearest Centroid Classifier (VALENCIA)

The taxonomy of cervical microbial communities was sorted into categories termed community state types (CSTs). This categorization collapses a taxonomic profile into a single categorical variable, allowing data exploration and statistical modeling (14). In addition, categorization allows unbiased analysis of small and sizable vaginal microbiota datasets, comparisons between datasets, and meta-analyses that combine multiple datasets. We used the data package at github.com/ravel-lab/VALENCIA to analyze the following standard parameters and categorize our cervicovaginal communities into different CST and sub-CST categories (14).

### Analyses of Microbial Community Structure and Diversity

For community-level analyses, we computed the pairwise Bray-Curtis distances (Beta diversity) which quantify the compositional dissimilarity between samples (29). We visualized bacterial community composition and structure differences using non-metric multidimensional scaling (NMDS). Alpha diversity measure of Chao1 (richness), and Shannon index (30) (diversity) were computed and plotted using R with packages: “phyloseq”, “vegan” and “ggplot2” (31–33). Bar Plots showing genus and species-level taxa were computed using a locally run version of QIIME2 (28).

The metadata variables included in our study were: 1) cervical disease, defined as the cytological phenotype of negative for intraepithelial lesion or malignancy (NILM), low-grade squamous intraepithelial lesion (LGSIL) and high-grade squamous intraepithelial lesion (HGSIL); 2) HPV risk, defined as HPV negative, HPV positive with co-infection (low and high-risk HPV simultaneously), and HPV positive high-risk (exclusively high-risk types); 3) cytokine ranks (estimated through quartile and tertile distribution of the fluorescence scores as detected by Luminex MAGPIX® system); and 4) community state types (CSTs).

### Statistical Analyses of the Microbiota Profiles

For beta diversity analyses, we tested if distances between sample groups were statistically significant using ANOSIM, a non-parametric statistical test (34). Using a Bray-Curtis distance table, we used this test to compare ranked beta diversity distances between the different variables. These tests were done using the script “qiime diversity beta-group-significance” for each specific test in QIIME2 (28) with the distance matrix as the input file and 999 permutations (34). Statistical analyses for alpha diversity were done using the script “qiime diversity alpha-group-significance” in QIIME2 (28), which uses a non-parametric t-test with Monte Carlo permutations. Additionally, to ascertain if there are statistically significant differences between the ASV abundance in the different sample groups, we used linear discriminant analysis (LDA) effect size (LEfSe) (35) through R, using MicrobiomeAnalyst (36). Significant ASVs were determined considering a p-value of < 0.05 and LDA scores of 1.5 for all cytokine groups. Cytokine fluorescence values (as logarithms) were associated to differentially abundant levels of *L. crispatus*, *L. iners*, *Sneathia*, *Gardnerella*, *Atopobium* and *Prevotella* using a linear mixed model (LMM) in R (ggplot and reshape2 libraries), correcting for age and body mass index (BMI). Differential abundance analysis was performed using amplicon sequence variants corresponding to *L. crispatus*, *L. iners*, *Sneathia*, *Atopobium*, *Gardnerella* and *Prevotella*. They were evaluated for their differential presence across the cytokine groups. Initially, a summary of each one of the ASVs of interest was used to perform groupings due to abundance. The groupings were separated as follows ASV 0-50 counts = very low abundance; ASV count 51-500 = low abundance; and >500 = high abundance.

## Results

### Population description and Community State Types (CSTs) prevalence

A total of 91 women were included in the study. The mean age was 39 years old. Women had an average pH value of 5.49. The distribution of the groups of samples according to cervical neoplasia severity included Negative for Intraepithelial Lesion or Malignancy (NILM) (N=56) Low-Grade Squamous Intraepithelial Lesion (LGSIL) (N=18), and High Grade Squamous Intraepithelial Lesion (HGSIL) (N=17), with differential HPV status and cytokine ranks (Table 1, S1 Table). HPV infections had a prevalence of 67.0% (including infections with any HPV type) with a higher prevalence of HPV+ among women with LGSIL (94.4%) and HSIL lesions (88.2%) (Table 1). As expected, HR-HPV prevalence was higher in HGSIL (82.4%) compared to in LGSIL (50.0%) and negative cervical lesions (39.3%) (p=0.015). Participants with HPV co-infection were primarily observed in participants with low-grade lesions (44.4%) (p=0.040), and less abundant in the group with high-grade lesions (5.9%). There were no significant differences in the distribution of the BMI groups across cervical disease, with equal proportions of normal weight, overweight and obese women (Table1). Across all women, cervical lesion prevalence was 38.5% (including LGSIL and HGSIL), Individually, LGSIL and HGSIL comprised 19.8% and 18.7% of the samples, respectively. CST1 mainly was present in 16% of the NILM cases and 22% LGSIL while not being present in HGSIL. In contrast, CST IV-B was only identified on participants with high-grade lesions (HGSIL) (p=0.025) (Table 1). Prevalence analysis of CSTs per levels (high and low) of cytokine groups (pro-inflammatory, traffic-associated, and anti-inflammatory) revealed that CST IV-C is the more prevalent in all cytokines (S1 Fig., S2 Table).

**Table 1.**
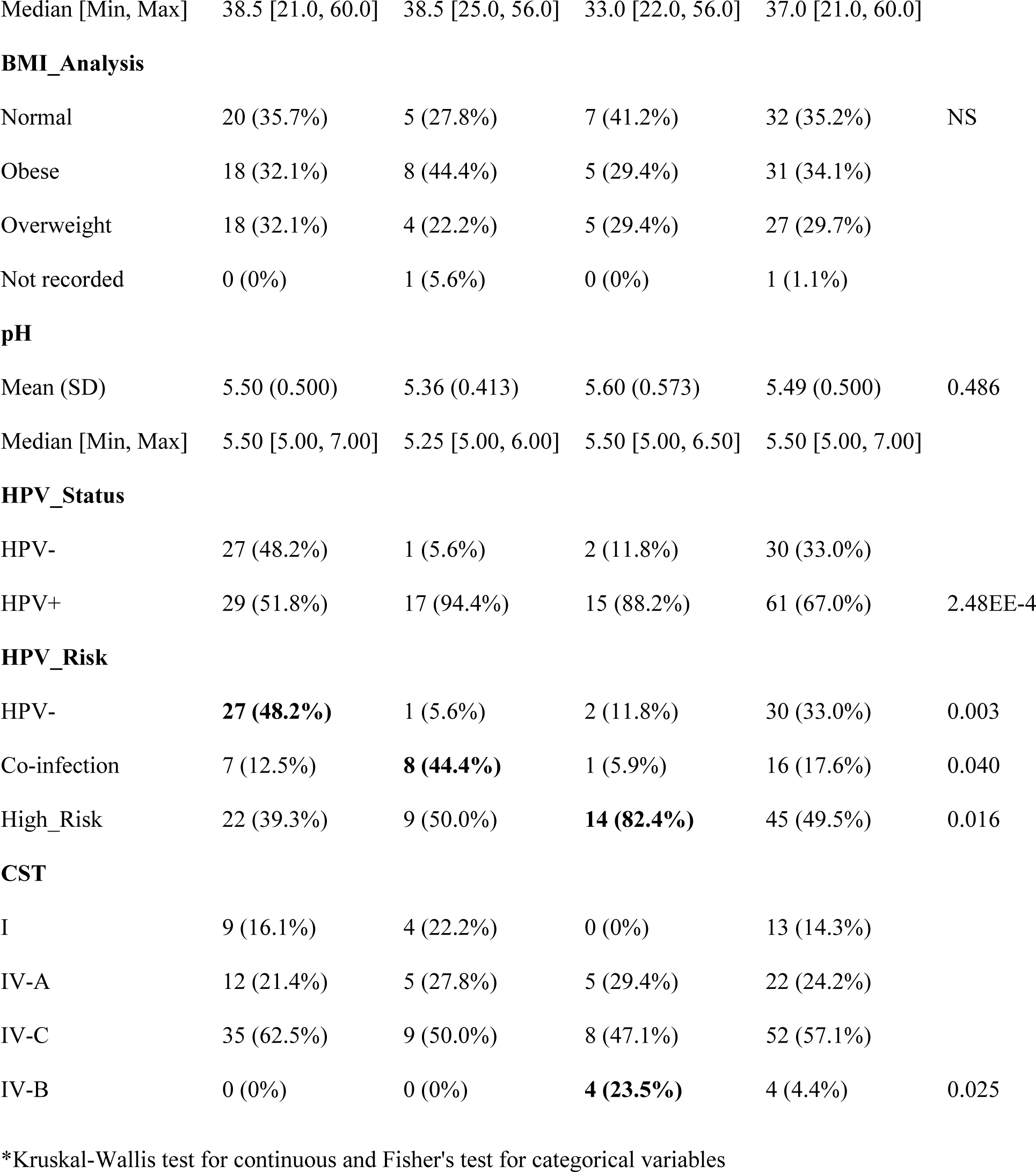
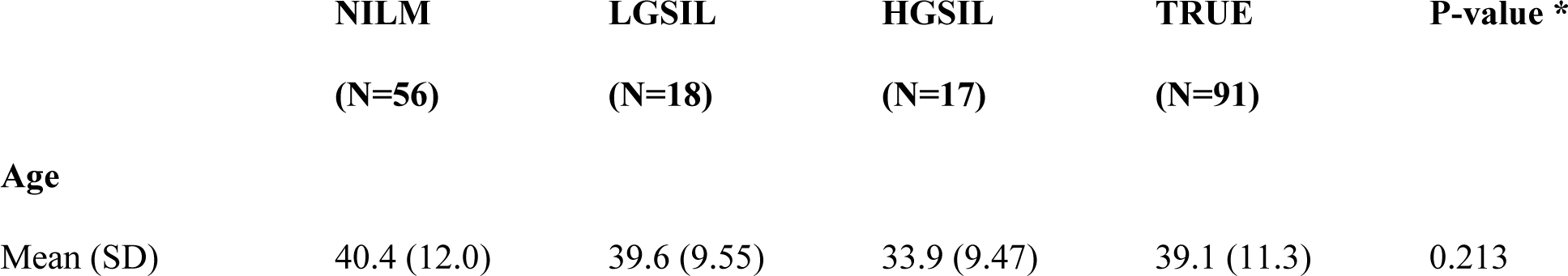
Characteristics of the population.

|  | **NILM**<br>**(N=56)** | **LGSIL**<br>**(N=18)** | **HGSIL**<br>**(N=17)** | **TRUE**<br>**(N=91)** | **P-value \*** |
| --- | --- | --- | --- | --- | --- |
| **Age** |  |  |  |  |  |
| Mean (SD) | 40.4 (12.0) | 39.6 (9.55) | 33.9 (9.47) | 39.1 (11.3) | 0.213 |
| Median [Min, Max] | 38.5 [21.0, 60.0] | 38.5 [25.0, 56.0] | 33.0 [22.0, 56.0] | 37.0 [21.0, 60.0] |  |
| BMI\_Analysis |  |  |  |  |  |
| Normal | 20 (35.7%) | 5 (27.8%) | 7 (41.2%) | 32 (35.2%) | NS |
| Obese | 18 (32.1%) | 8 (44.4%) | 5 (29.4%) | 31 (34.1%) |  |
| Overweight | 18 (32.1%) | 4 (22.2%) | 5 (29.4%) | 27 (29.7%) |  |
| Not recorded | 0 (0%) | 1 (5.6%) | 0 (0%) | 1 (1.1%) |  |
| pH |  |  |  |  |  |
| Mean (SD) | 5.50 (0.500) | 5.36 (0.413) | 5.60 (0.573) | 5.49 (0.500) | 0.486 |
| Median [Min, Max] | 5.50 [5.00, 7.00] | 5.25 [5.00, 6.00] | 5.50 [5.00, 6.50] | 5.50 [5.00, 7.00] |  |
| HPV\_Status |  |  |  |  |  |
| HPV- | 27 (48.2%) | 1 (5.6%) | 2 (11.8%) | 30 (33.0%) |  |
| HPV+ | 29 (51.8%) | 17 (94.4%) | 15 (88.2%) | 61 (67.0%) | 2.48EE-4 |
| HPV\_Risk |  |  |  |  |  |
| HPV- | **27 (48.2%)** | 1 (5.6%) | 2 (11.8%) | 30 (33.0%) | 0.003 |
| Co-infection | 7 (12.5%) | **8 (44.4%)** | 1 (5.9%) | 16 (17.6%) | 0.040 |
| High_Risk | 22 (39.3%) | 9 (50.0%) | **14 (82.4%)** | 45 (49.5%) | 0.016 |
| CST |  |  |  |  |  |
| I | 9 (16.1%) | 4 (22.2%) | 0 (0%) | 13 (14.3%) |  |
| IV-A | 12 (21.4%) | 5 (27.8%) | 5 (29.4%) | 22 (24.2%) |  |
| IV-C | 35 (62.5%) | 9 (50.0%) | 8 (47.1%) | 52 (57.1%) |  |
| IV-B | 0 (0%) | 0 (0%) | **4 (23.5%)** | 4 (4.4%) | 0.025 |

\*Kruskal-Wallis test for continuous and Fisher's test for categorical variables

### Cytokine changes according to high-grade lesions and community state types (CSTs)

There were significant differences in fluorescence between pro-inflammatory cytokines IL-1β and IFNγ among women with high grade disease, compared to healthy participants. There was significantly higher fluorescence values between HGSIL and (NILM / HPV-p=0.008) or with participants with low-grade disease (p=0.015) (Fig. 1 A). Although there were no significant differences among groups when evaluating traffic-associated cytokines (Fig. 1B) or anti-inflammatory cytokines (Fig. 1C), there was a tendency for higher fluorescence levels in participants with any cervical disease (LGSIL and HGSIL). Although an analysis evaluating the cytokines according to HPV risk was done, no significant differences were identified (S2 Fig.).

**Fig. 1.**
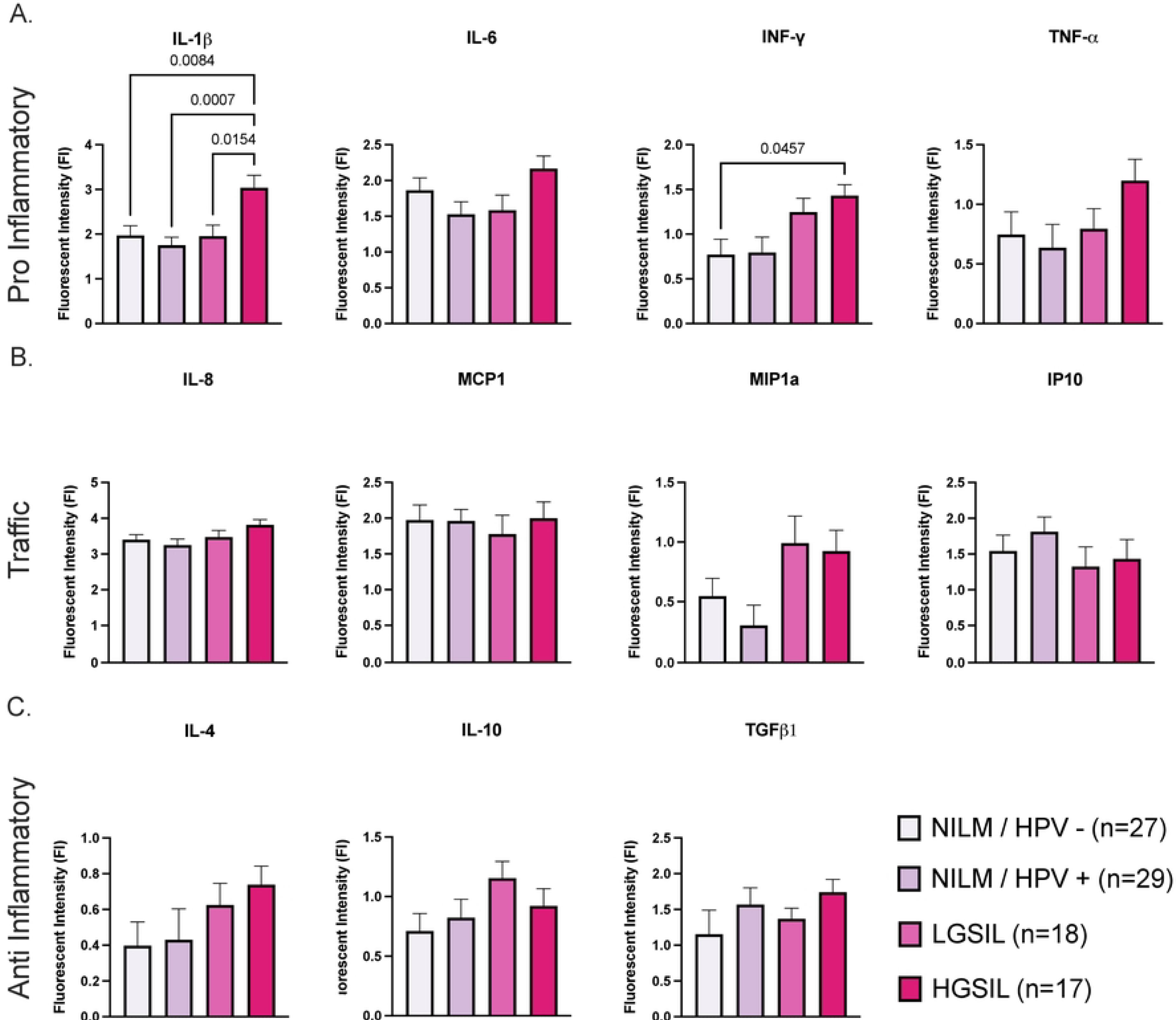
Cytokine Profiles grouped as pro inflammatory. **(A)** (IL-1β, TNFɑ, IFN𝞬, IL-6), anti-inflammatory **(B)** (IL-4, IL-10, TGFβ1) and traffic-associated cytokines**(C)** (IL-8, MIP1a, MCP1, IP10) cytokines evaluated through LUMINEX and plotted according to cervical disease. Fluorescence Intensity values were used to compute multiple comparison analysis using ordinary one-way ANOVA with Tukey’s multiple comparisons test. Results were depicted in boxplots for cervical disease. Significant differences are highlighted by brackets and corresponding p-values.

Evaluation of cytokines according to CSTs revealed significant differences in IL-1β, IL-6, MIP1a and IP10 concentrations. CST IV-B was found to be associated with a significantly higher concentration of IL-1β (p=1.0EE-4), IL-6 (p=0.050) and MIP1a (p=0.008) when compared to *Lactobacillus*-dominated CST I. (Fig. 2). In contrast, IP10 was significantly higher in CST I compared to CST IV-A (p=0.031) (Fig. 2).

**Fig. 2.**
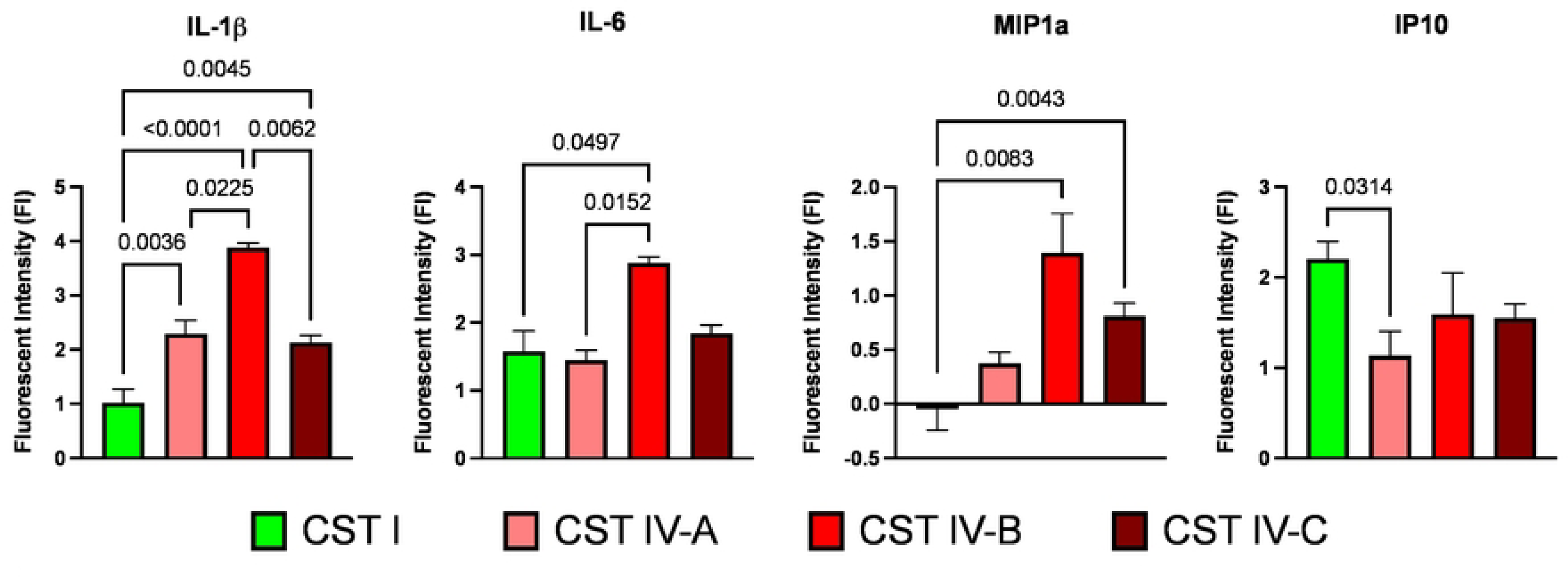
Cytokine Profiles of IL-1β, IL-6, MIP1a and IP10 cytokines evaluated through LUMINEX, plotted according to CST classification. Fluorescence Intensity values were used to compute multiple comparison analyses using ordinary one-way ANOVA with Tukey’s multiple comparisons tests. Results are depicted in boxplots for cervical disease, and brackets and corresponding p-values highlight significant differences.

### HPV infection and cervical disease are associated with altered bacterial community composition

A total of 2,067,074 good-quality 16S rRNA sequence reads and 8,167 OTUs were analyzed, with a rarefaction level of 7,400 reads per sample. Our samples mainly included reproductive-age women (n = 73) and women in menopause (n = 18). To evaluate whether changes in cervicovaginal microbiota are associated according to the women’s physiological state, we analyzed beta and alpha diversity between these categories. We found no significant differences neither in beta (Bray-Curtis) (S3 Fig. A) nor in alpha diversity (Shannon) (S3 Fig. B). The microbiota composition did not reveal any unique taxa associated with any of the evaluated groups (S3 Fig. C). Additionally, we evaluated the cytokine profiles associated to menopause state (S3 Fig. D, E and F). We found no significant differences among menopause and non-menopause women according to the cytokine profiles. All the associations of cytokines with specific taxa were corrected for age.

We evaluated the differences in the microbiota according to cervical disease and HPV risk. Beta diversity analyses considering cervical disease, revealed that bacterial community structure was significantly different according to cervical disease severity (LGSIL p=0.026) (HGSIL p=0.003) when compared to healthy controls (NILM/HPV-) (Fig. 3A). As for HPV risk, community structure was found to be significantly altered due to HR-HPV infection (p=0.042) and HPV co-infections (p=0.004) compared to HPV negative samples (Fig. 3D, S3 Table). Alpha diversity (Shannon) increased concomitantly to the severity of the epithelial lesions, including significant differences between NILM and HGSIL (p=0.003), regardless of HPV infection (Fig. 3B, S4 Table). In contrast, alpha diversity increased in participants with HPV co-infections as compared to HPV negatives (p=0.032) (Fig. 3E, S4 Table).

**Fig. 3.**
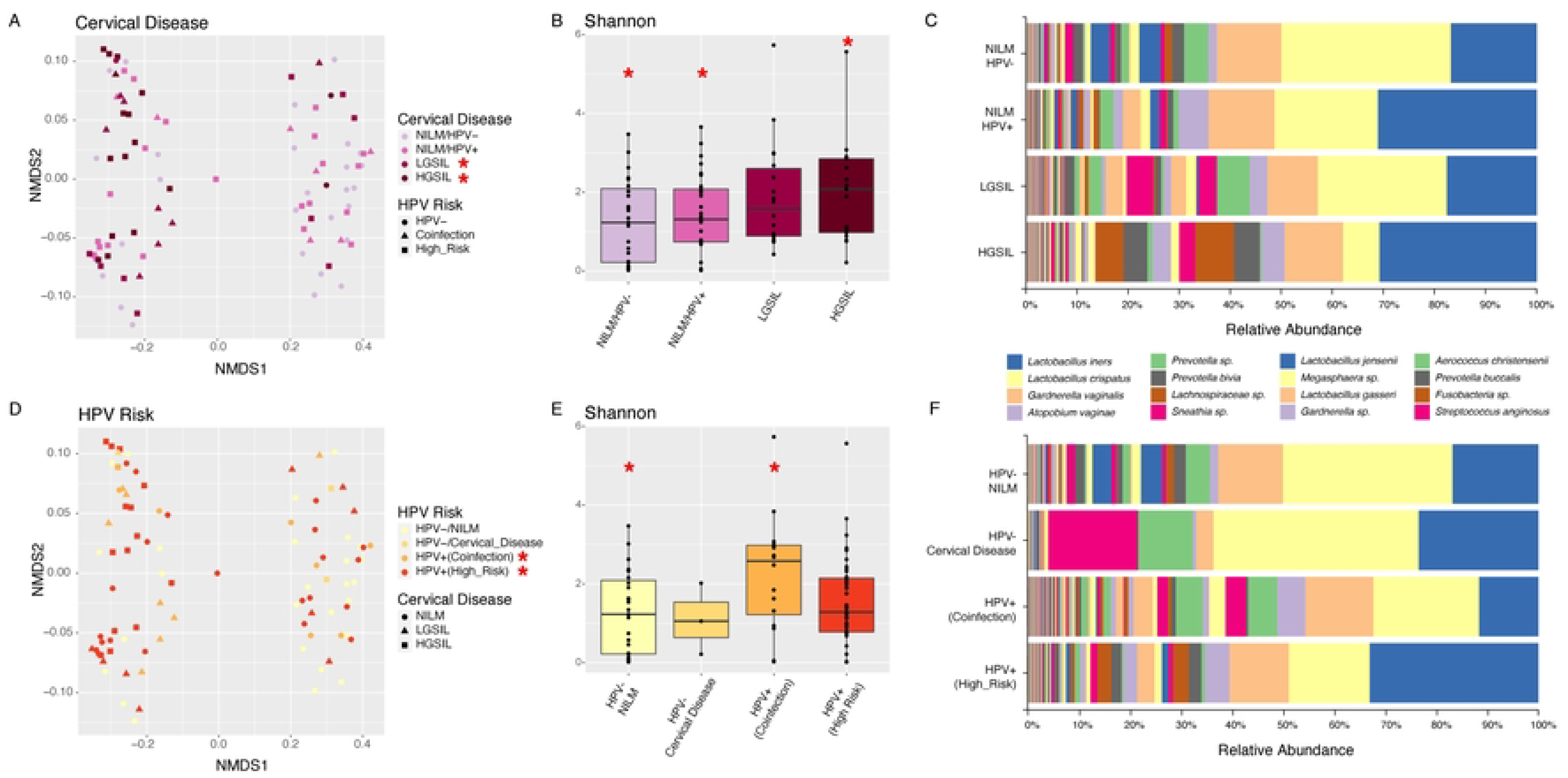
Diversity analyses according to cervical disease and HPV risk categories. Bray-Curtis analysis represented by Non-metric multidimensional scaling (NMDS) using cervical disease and HPV risk as metadata categories (A, D). Alpha diversity (Shannon) was calculated and depicted in Figs B and E. Bar Plots depicting relative abundance of bacteria at the species level for cervical disease (C) and HPV risk (F). Red asterisks mean significant differences among groups (p<0.05).

The microbiota composition between NILM and LGSIL participants was not different and was dominated by *L. crispatus* and *L. inners*. Participants with HGSIL had a change in *Lactobacillus* distribution, being dominated by *L. inners* and other taxa associated with dysbiosis, such as *Lachnospiraceae sp., P. bivia, A. vaginae* and *G. vaginalis* (Fig. 3C). Bacterial composition changed dramatically according to HPV Risk, favoring a predominance of *L. crispatus* in HPV negative women, and *L. iners* in high-risk participants (Fig. 3F). Participants with HPV co-infections had a highly diverse taxonomic profile with dysbiosis associated taxa including, *Prevotella sp*., *Megasphaera sp*., *Sneathia*, *Aerococcus christinsenii* and *Streptococcus anginosus* (Fig. 3F).

As identified by linear discriminant analysis (LDA) effect size (LEfSe), putative microbial biomarkers detected *Parabacteroides* and *Akkermansia* in participants with HGSIL. As for HPV risk, we found that *Sneathia* was more abundant in HPV-negative participants with cervical disease. In addition, for HPV co-infections, we identified *Megasphaera*, *Parabacteroides*, *Akkermansia* and *Bacteroides* as significantly differentiated taxa (Fig. 4).

**Fig. 4.**
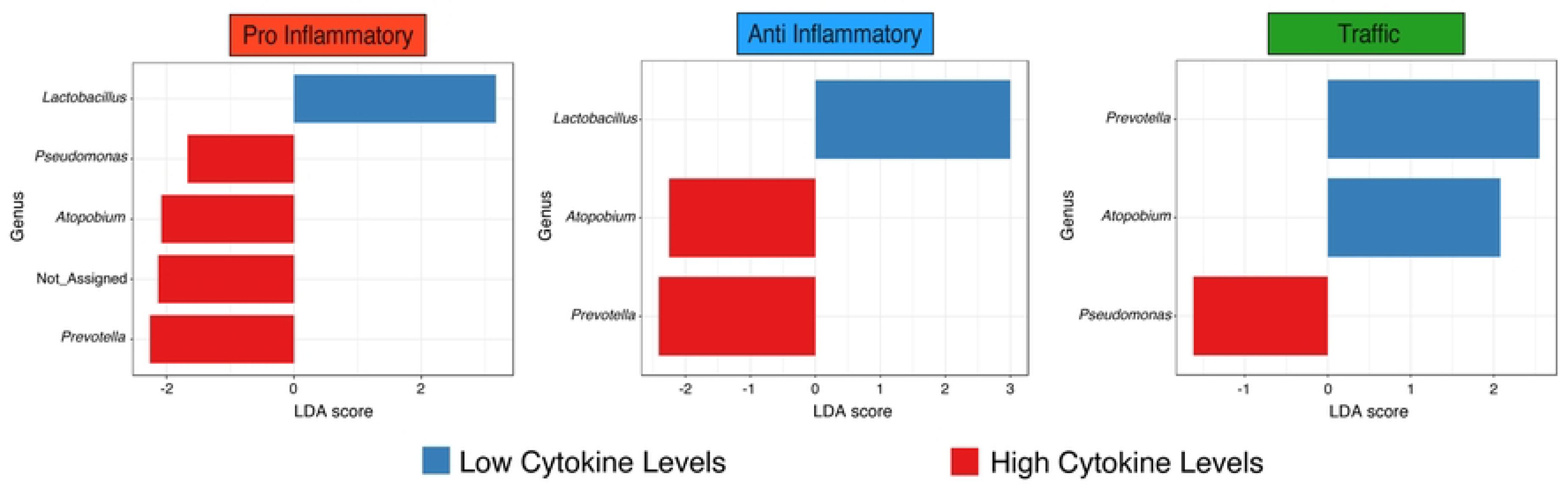
Linear discriminant analysis (LDA) effect size (LEfSe) for cervical disease and HPV risk at the Genus level. Cutoff was 1.5 LDA score and a p value of 0.05. Log-level changes in LDA score are displayed on the x axis.

### Association of bacterial taxa with cytokine levels

In a balanced vaginal tract, *Lactobacillus* species impede the colonization of other bacteria that would otherwise cause infections. *L. Crispatus* produce lactic acid, bacteriocins and H_2_O_2_ acting as a chemical barrier (Valenti et al. 2018). Very low levels of *L. crispatus*, a key vaginal bacterium in eubiosis, were significantly associated to high concentrations of pro-inflammatory cytokines IL-1β (p=2.16E-6) and IFNγ (p=0.013) (Fig. 5). High levels of *L. iners* were significantly associated with high levels of IL-6, also a pro-inflammatory cytokine (p=0.019), suggesting that this species of *Lactobacillus* might stimulate IL6 production (Fig. 5). On the contrary, low levels of *L. iners* correlate with lower expression of IFNγ (p=0.039) (Fig. 5). Very low levels of *Atopobium,* a mostly pathogenic bacteria were significantly associated with low levels of IL-1β (p=0.002). As for *Sneathia,* another dysbiosis associated taxa, detected at low levels, had correlation with higher levels of IFNγ (p=7.00E-6), TNFα (p=7.00E-4) TNFα (p=0.010) (Fig. 5). Absence, or very low levels anaerobic *Gardnerella,* were associated with lower fluorescence values of IL-1β (p=1.00E-6) (Fig. 5). Very low abundance *of Prevotella* correlated with lower concentrations of IL-1β in our samples (p=3.00E-6) (Fig. 5).

**Fig. 5.**
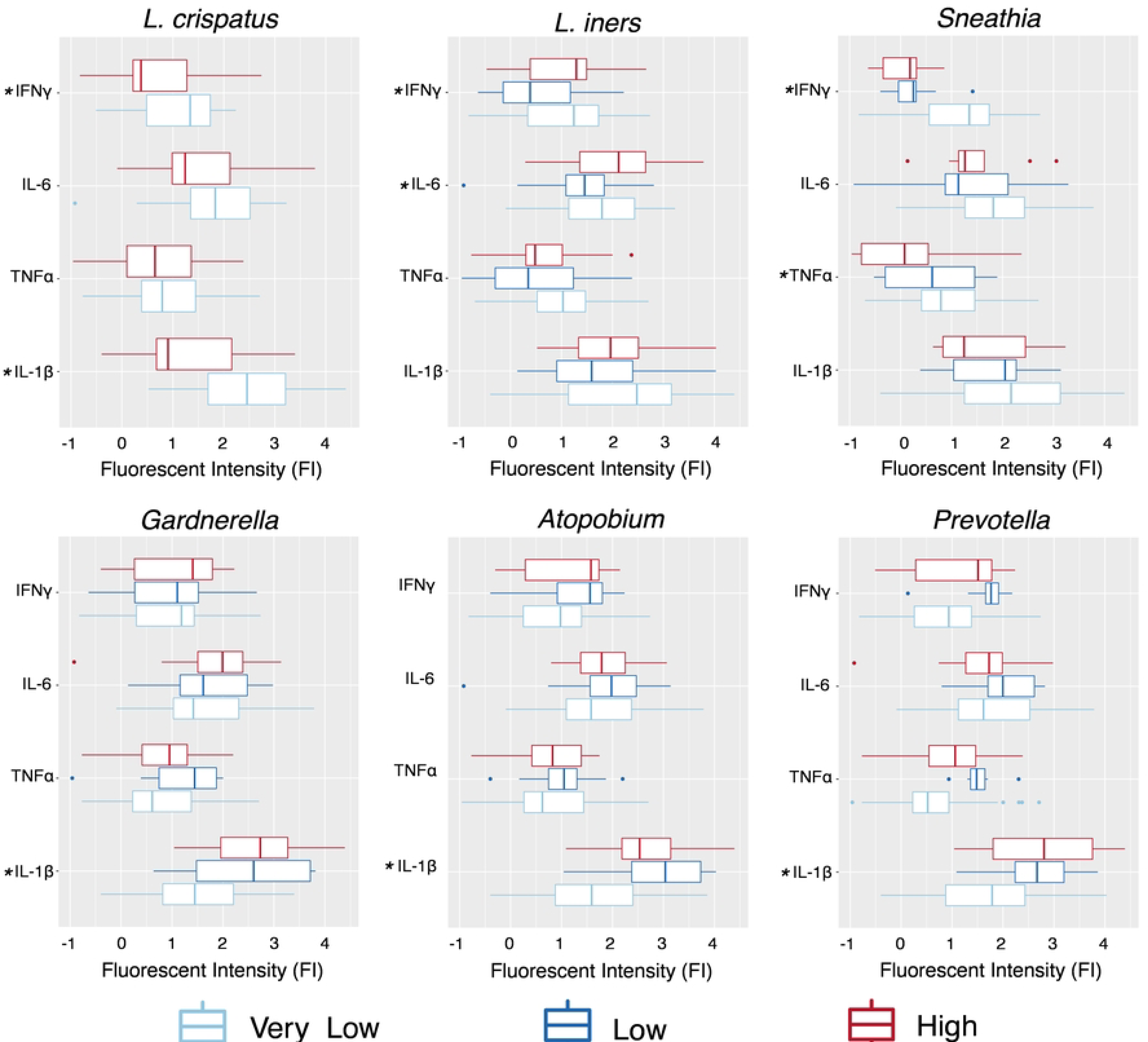
Association of key vaginal bacteria with levels of pro-inflammatory cytokines. Cytokine logarithm fluorescence values were associated to differentially abundant levels of *L. crispatus*, *L. iners*, *Sneathia*, *Gardnerella*, *Atopobium* and *Prevotella* using a linear mixed model (LMM), correcting for age and BMI. Differential abundance groupings were separated as follows ASV 0-50 counts = very low abundance; ASV count 51-500 = low abundance; and >500 = high abundance.

In beta diversity, axis 1 of the NMDS1 plot separated samples according to their cytokine levels (S4 Fig. A, D, G). When compared to healthy controls (NILM/HPV-) we found changes in microbial community structure significantly associated with high levels of pro-inflammatory cytokines regardless of HPV infection (NILM/HPV-, p=0.006; NILM/HPV+, p=0.001) and cervical disease severity (LGSIL, p=0.002; HGSIL, p=0.001) (S4 Fig. A, S3 Table). High levels of anti-inflammatory cytokines associated with changes in bacterial composition in similar patterns as pro-inflammatory cytokines, with significant differences regardless of HPV infection (NILM/HPV-, p=0.001; NILM/HPV+, p=0.001) and cervical disease severity (LGSIL, p=0.012; HGSIL, p=0.001) (S4 Fig. D, S3 Table). High levels of traffic-associated cytokines significantly altered bacterial composition regardless of cervical disease severity (LGSIL, p=0.012; High_HGSIL, p=0.001) (S4 Fig. G, S3 Table). Alpha diversity significantly increased with cytokine levels for both pro-inflammatory (NILM/HPV-, p=0.035; NILM/HPV+, p=0.002) (LGSIL, p=0.002; HGSIL p=0.001) and anti-inflammatory (NILM/HPV-, p=0.039; NILM/HPV+, p=0.005) (LGSIL, p=0.001; HGSIL, p=3.55E-4) cytokines regardless of HPV infection and cervical disease (S4 Fig. B, 4E). Traffic-associated cytokines did not have significant alpha diversity differences associated with cytokine concentrations (S4 Fig. H, S4 Table). Taxonomic abundance at species level showed that in the 3 cytokine groups, populations of *L. crispatus* decreases as cytokine levels and cervical disease severity increased. Participants with high levels of cytokines and high-grade cervical disease were dominated by *L. iners*, as *L. crispatus* nearly disappears. Additionally, this group of participants were dominated by *Prevotella sp*., *A. vaginae* and *G. vaginalis*. Simultaneously, *Lachnospiraceae sp.* was detected mainly on participants with High cytokine levels and HGSIL, confirming the dysbiosis typical of *Lactobacillus* depleted communities (S4 Fig. C, 4F and 4I). LefSe analysis (p=0.05 and 1.5 LDA score cutoffs) revealed an increase in *Lactobacillus* when pro-inflammatory and anti-inflammatory cytokines were low (Fig. 6). Low levels of traffic-associated cytokines presented anaerobic bacteria like *Prevotella* and *Atopobium* with higher abundance (Fig. 6). In contrast, with high cytokine levels in pro-inflammatory and anti-inflammatory cytokines, *Atopobium* and *Prevotella* populations decreased. *Pseudomonas* communities also were reduced in association with pro-inflammatory and traffic-associated cytokines (Fig. 6).

**Fig. 6.**
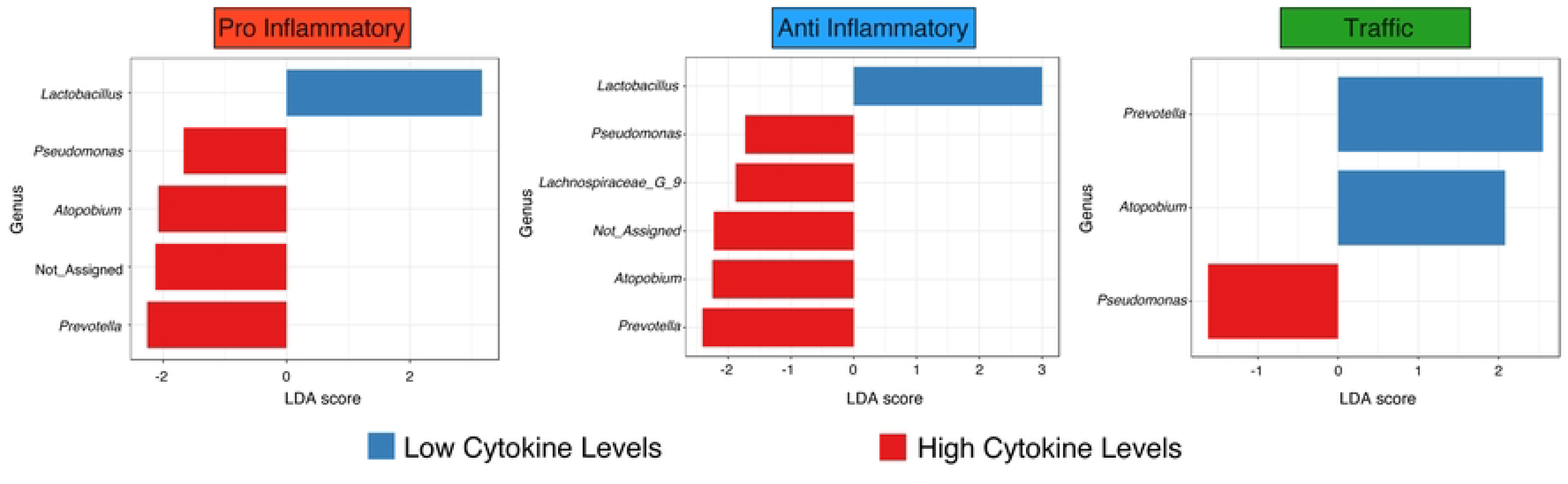
Linear discriminant analysis (LDA) effect size (LEfSe) for **(A)** pro-inflammatory (IL-1β, TNFɑ, IFN𝞬, IL-6) (LDA score 1.5), **(B)** anti-inflammatory (IL-4, IL-10, TGFβ1) (LDA score 2.25) and **(C)** traffic (IL-8, MIP1a, MCP1, IP10) (1.5 LDA score) cytokines evaluated considering low and high cytokine levels and a p-value cutoff of 0.05.

## Discussion

Cervical cancer (CC) is a complex disease where the cervical environment is modified, including cytokine expression that leads to a local immunosuppression state (18). Approximately 15 high-risk HPV strains cause most cervical cancers; however the detection of HPV has a low positive predictive value for invasive cervical cancer, as few HPV+ women progress to High-Grade Squamous Intraepithelial Lesions (HGSIL) (6,37). Inflammation and the microbiome could be the missing link to understanding the actual cellular mechanisms leading to dysplasia and cancer. Indeed, to provide an accurate diagnosis and prognosis of cervical disease progression, screening tests have limited sensitivity for histopathology and cytology (38). Our sensitive HPV genotyping method demonstrated that HPV prevalence in this cohort was 67.0% (encompassing infections with any HPV type) and that co-infections with any HPV serotype were primarily present in low-grade lesions (44.4%) and exclusively high-risk types were more prevalent in HGSIL (82.3%), a prevalence previously reported by our team (11) and Hispanic and Amerindian peoples (39). This higher prevalence exceeds those reported for Costa Ricans (40), or women in Surinam (41), Nigeria (42), Turkey (43) or Italy (44). This can be attributed to the fact that these women are recruited in colposcopy clinics and are therefore not representative of the Puerto Rican population.

Modifying the host innate immune system is a well-known feature of numerous malignancies, including HPV-driven cancers (45). HPV interferes with the cytokines, Toll-like receptors (TLRs), and the overall inflammatory process, occurring in the early stages of the immune response (46,47). Together, cytokines and TLRs identify HPV and encourage immunological signalings such as cell development and differentiation (45,47). In cervical HPV infections, an impaired immune response to the virus may lead to cervical disease progression (48). Overall, we found that any HPV infection elevated all cytokine levels but not all cytokines were significant. This suggests that HPV infections alone cannot drive cervical dysplasia progression to cancer (49). Most HR-HPV-infected women do not develop CC because an effective immune response controls the infection and therefore, malignant transformation is halted (50). Additionally, HPV uses several strategies to evade the immune system (Miura et al., 2010), such as reducing the amount of IFNγ, altering the activation of antiviral and antitumor immune cells such as NK cells and T cells with antiviral and antitumor activity (51). In addition to this, an imbalance of the mucosal microbiome, such as a loss in protective *Lactobacillu*s and physicochemical changes, causes alterations of the vaginal mucosa and the cervical epithelium, exercising a pattern of selective pressure on the microbiota (52).

Several studies have already established background data on how the microbiome is modified by HPV infections, stating that HPV infections alone are not enough to drive significant changes that translate into epithelial modifications to cancer (11,53). Nonetheless, other studies suggested changes in the composition of the vaginal microbiota associated with human papillomavirus infection (54–56). Our results show greater microbiota diversity and a lower abundance of lactobacilli among HPV-positive women, which is congruent with previous studies (J. E. Lee et al. 2013; Oh et al. 2015; Reimers et al. 2016, Godoy-Vitorino et al. 2018). It is still unclear why some HR-HPV infections clear while others persist, causing dysplasia and even cervical cancer. Therefore, since only persistent HR-HPV infection has been associated with immune response and vaginal dysbiosis, molecular differences between transient and persistent HR-HPV infection may hold information to explain this phenomenon (37).

Concerning cervical lesions, we found a strong microbiome association with the cervical phenotype (lesion severity), with a significantly higher diversity value (Shannon) in HGSIL compared to NILM/HPV- and NILM HPV+ participants, but not substantially different from LGSIL. Our findings coincide with most studies suggesting an association between *L. iners* and HGSIL (60,61). *Lactobacillus* produces have several mechanisms that function as a defensive barrier for invasive bacteria, such as, the production of significant amounts of bacteriocins that contributes to colonization the use of available glycogen to produce lactic acid, lowering vaginal pH, responsible of lysing bacteria other than *Lactobacillus* (62–65)(Amir et al. 2020; Cruickshank 1934; O’Hanlon et al. 2013; Zhou et al. 2019). The loss of this defensive strategy due to *Lactobacillus* community reduction, explains the increase of *Atopobium vaginae*, *Prevotella* spp., and *Gardnerella* spp.

Our study confirms cervicovaginal microbial dysbiosis in high-risk HPV and cervical disease severity associated with pro-inflammatory cytokines, validating other studies in other cohorts (18,66). IL-1β and IFNγ were significantly more abundant in high-grade disease participants, associated with a loss in *Lactobacillus* validating previous studies where high concentrations of pro-inflammatory cytokines were detected in women with cervical neoplasia (67–69). The rise in IL-1β can be attributed to cellular mechanisms such as tumorigenesis, angiogenesis (70) or metastasis (71); as well as increased risk of CC in women with high grade cervical lesions (72) associated with either polymorphism (73), reactive oxygen species (ROS), COX2 or NADPH oxidase (74). We found higher levels of expression of IL-6 associated with high levels of *L. iners.* IL-6 is essential for the growth, metastasis, and initiation of cancer; affecting tumor growth, differentiation, and the host immune defense mechanism (75). Many immune and nonimmune cells, such as T cells, monocytes, macrophages, dendritic cells, and some cancer cells -such as CC cells-produce IL-6 (76,77). IL-6 induces CC growth by vascular endothelial growth factor (VEGF)-dependent angiogenesis (78). In addition, higher expression of IL-6 was shown in cancerous tissues than in adjacent non-cancer tissues in early-stage CC patients, confirming its association to cancer development (79). The less protective Lactobacilli (*L. iners)* may be stimulating the production of these mediators, this could be due in part to their heterogeneous genome, that unlike the protective counterparts, do not produce as much lactic acid or hydrogen peroxide (80). Although the slight increase in IL-10 among LGSIL is not significant, we hypothesize that IL-10 may increase in parallel with the development of intraepithelial lesion as it does in cancer, promoting the expression of HPV oncoproteins (Berti et al. 2017).

Taxonomic profiles of cervical microbial communities were sorted into community state types (CSTs), allowing a valuable categorization for statistical modeling and comparisons between datasets (13,14). The vaginal microbiota in this Hispanic women cohort of women living in Puerto Rico was dominated by CST IV-C (57.1%, non-*Lactobacillus* profile) and CST IV-A (24.1%, *G. vaginalis* moderate), which confirms previous work of our lab (11), and suggests similarities to other US Hispanics (13) and Venezuelans (81). Even when CST IV-C dominated all three categories of cervical cytology (NILM, LGSIL, HGSIL), but was more predominant in healthy patients (NILM) (62.5%). CST IV-C is characterized by a low relative abundance of *Lactobacillus spp*., *G. vaginalis*, and *A. vaginae*, while instead is characterized by the abundance of a diverse array of facultative and strictly anaerobic bacteria, thus revealing that Hispanic women living in PR are in constant dysbiosis with a microbiome dominated by anaerobic non-lactobacillus communities. Overall, the high prevalence of CST-IV across all samples reflects a cervical microbial environment with low stability and less likelihood to return to its equilibrium state after a perturbation, which increases the risk of developing adverse health outcomes (82,83). In Hispanics living in Puerto Rico, the typical healthy vaginal microbial profile seems to be already diverse. It does not correspond to that of US Caucasians, mostly dominated by CST-I, being more like that of other US Hispanics, which are more prone to vaginal dysbiosis regulation and inflammatory conditions (Ravel et al. 2011). CST-IV represents a highly diverse bacterial profile and has been associated with high pro-inflammatory cytokine levels, which damage the endocervix’s columnar epithelial barrier, exposing it to colonization to other microbial or pathogenic agents (12,84).

Microbiome analysis revealed that high levels of all three groups of cytokines (anti-inflammatory, pro-inflammatory, and traffic-associated) were related to significant bacterial community composition changes and high diversity levels (Shannon). Low levels of *Sneathia spp*. are associated with higher concentrations of IL-1β and TNFα, this taxa that is part of the normal microbiota of the genitourinary tracts of men and women is now considered a possible pathogen of the female reproductive tract (85), *Sneathia spp*. are also regarded as good biomarkers for bacterial vaginosis (86), a classical dysbiosis associated with abnormalities in cervical smears (87). Previous studies have described that *Lactobacillus* can prevent the colonization of exogenous pathogens by producing lactic acid, bacteriocins and reactive oxygen species (ROS) (13,88,89). Lactobacilli, promote homeostasis in the cervicovaginal environment and are associated with low levels of pro-inflammatory and anti-inflammatory cytokines. In contrast, low concentrations of traffic-associated-related cytokines are associated with an increase in *Prevotella* and *Atopobium*, suggesting that there is little induction of immune cell tissue migration to combat these bacteria. Previous studies reported that *Atopobium vaginae* could trigger an innate immune response involving IL-6 and IL-8 using an in vitro model of bacterial vaginosis (90), and this could be happening *in vivo*. Also, a study suggested that *Prevotella* may provide nutrients such as ammonia and amino acids to other microbial community members such as *Gardnerella* and *Peptostreptococcus* (91), assuming a metabolic hub for vaginal microbiota and being a key player in vaginal dysbiosis (92). *Prevotella* is therefore critical for maintaining a dysbiotic ecosystem in the cervix.

Cervical cancer is a complex multifactorial disease, and further research, including time-series analyses of longitudinal sampling, is needed to improve our understanding of its etiology. Our study evidence that high concentrations of all three cytokine groups (inflammatory, anti-inflammatory, and traffic-associated) were associated with cervical dysbiosis, which occurs through dramatic losses of protective *Lactobacilli* and the presence of diverse anaerobic taxa. The cervical microbiota in Hispanic women living in Puerto Rico is highly diverse and dominated by CSTs III and IV. Further changes of the microbiota with HR-HPV and inflammation reveal that the joint host-microbe interaction analyses via cytokine signaling and microbiota in precancerous lesions have great translational potential.

## Data Availability

The 16S-rRNA reads and corresponding sample metadata was deposited in QIITA study ID 12871 and the raw sequences are available in the European Nucleotide Archive ENA Project under accession number EBI: ERP136546. https://www.ebi.ac.uk/ena/browser/view/PRJEB51893

## Acknowledgments

We would like to acknowledge Dr. Robert D. Burk, Dr. Mykhaylo Usyk for kindly providing the curated extended GreenGenes database we cite in the methods section, which integrates the GreenGenes database, the Human Oral Microbiome Database (HOMD) and cervicovaginal microbiome 16S reference sequences retrieved from NCBI. Also, we want to thank Dr. Michael T. France for his help in adapting our taxonomy for the VALENCIA classification. We thank all the medical personnel from the gynecology clinics who participated in the screening program and sample collection and the students who helped with metadata curation. We offer special thanks to the participants for contributing to our study.

## Supporting information

**S1 Fig. CST prevalence distribution among cytokine levels and groups.** CSTs according to category groups were used to compute Fisher’s test using pairwise analysis to compare between groups. Results were depicted in boxplots for cervical disease. Significant differences are highlighted by brackets and corresponding p-values.

**S2 Fig. Cytokine Profiles** of (A) pro inflammatory (IL-1β, TNFɑ, IFN𝞬, IL-6), (B) anti-inflammatory (IL-4, IL-10, TGFβ1) and (C) traffic (IL-8, MIP1a, MCP1, IP10) cytokines evaluated through LUMINEX, plotted according to HPV risk. Fluorescence Intensity values were used to compute multiple comparison analysis using ordinary one-way ANOVA with Tukey’s multiple comparisons test. Results were depicted in boxplots for cervical disease. Significant differences are highlighted by brackets and corresponding p-values.

**S3 Fig. Diversity and cytokine profile analyses comparing samples, according to menopause status.** Beta and alpha diversity analyses are represented by non-metric multidimensional scaling (NMDS) (A) and Shannon index boxplots (B). Relative abundance of bacteria at the species level is shown in a relative abundance bar plot (C). Cytokine Profiles of pro inflammatory (D), anti-inflammatory (E) and traffic (F) cytokines, plotted according to HPV risk. Fluorescence Intensity values were used to compute multiple comparison analysis using ordinary one-way ANOVA with Tukey’s multiple comparisons test. Significant differences are highlighted by brackets and corresponding p-values.

**S4 Fig. Diversity analyses comparing samples, according to inflammatory (IL-1β, TNFa, IFNg, IL-6), anti-inflammatory (IL-4, IL-10, TGFβ1), and trafficking (IL-8, MIP1a, MCP1, IP10) cytokines.** Bray-Curtis analysis represented by Non-metric multidimensional scaling (NMDS) using pro inflammatory (A), anti-inflammatory (D) and trafficking cytokines (G) as metadata categories. Alpha diversity (Shannon) was calculated and depicted in Figures B, E and H. Bar Plots showing relative abundance of bacteria at the species level for cytokine groups (C, F, I).

**S1 Table. Study population characteristics (n=91) described by cervical disease status, including HPV risk, community state types, and cytokine ranks.**

**S2 Table. Pairwise comparisons using Pairwise comparison of proportions (Fisher) of the cytokine levels. P values were adjusted using Bonferroni corrections.**

**S3 Table. Non-parametric analyses using ranked beta diversity distances between different group depths found in the mapping file, based on the Bray-Curtis table.**

**S4 Table. Non-parametric t-test comparing observed richness between the different metadata categories**

## References

1. Mitchell C. PAHO/WHO | Cervical cancer is the third most common cancer among women in Latin America and the Caribbean, but it can be prevented [Internet]. Pan American Health Organization / World Health Organization. 2019 [cited 2022 May 17]. Available from: https://www3.paho.org/hq/index.php?option=com_content&view=article&id=14947:cervical-cancer-is-the-third-most-common-cancer-among-women-in-latin-america-and-the-caribbean-but-it-can-be-prevented&Itemid=1926&lang=en

2. Glasgow L, Lewis R, Charles S. The cancer epidemic in the Caribbean region: Further opportunities to reverse the disease trend. The Lancet Regional Health - Americas. 2022 Sep;13:100295.

3. Doorbar J, Egawa N, Griffin H, Kranjec C, Murakami I. Human papillomavirus molecular biology and disease association. Rev Med Virol. 2015 Mar;25 Suppl 1:2–23.

4. Lehoux M, D’Abramo CM, Archambault J. Molecular mechanisms of human papillomavirus-induced carcinogenesis. Public Health Genomics. 2009;12(5–6):268–80.

5. Pradhan SR, Mahata S, Ghosh D, Sahoo PK, Sarkar S, Pal R, et al. Human Papillomavirus Infections in Pregnant Women and Its Impact on Pregnancy Outcomes: Possible Mechanism of Self-Clearance [Internet]. Human Papillomavirus. IntechOpen; 2020 [cited 2022 May 17]. Available from: https://www.intechopen.com/chapters/70423

6. Beata S, Dariusz S, Marianna M, Hanna R, Zbigniew K, Luiza W, et al. The Role of Human Papillomavirus in Cervical Cancer. Int J Cancer Clin Res [Internet]. 2019 Sep 28 [cited 2022 May 17];6(5). Available from: https://www.clinmedjournals.org/articles/ijccr/international-journal-of-cancer-and-clinical-research-ijccr-6-125.php?jid=ijccr

7. Okunade KS. Human papillomavirus and cervical cancer. Journal of Obstetrics and Gynaecology. 2020 Jul 3;40(5):602–8.

8. Conesa-Zamora P, Ortiz-Reina S, Moya-Biosca J, Doménech-Peris A, Orantes-Casado FJ, Pérez-Guillermo M, et al. Genotype distribution of human papillomavirus (HPV) and co-infections in cervical cytologic specimens from two outpatient gynecological clinics in a region of southeast Spain. BMC Infect Dis. 2009 Aug 10;9:124.

9. Norenhag J, Du J, Olovsson M, Verstraelen H, Engstrand L, Brusselaers N. The vaginal microbiota, human papillomavirus and cervical dysplasia: a systematic review and network meta-analysis. BJOG. 2020 Jan;127(2):171–80.

10. Pfeiffer JK. Host response: Microbiota prime antiviral response. Nat Microbiol. 2016 Jan 27;1:15029.

11. Godoy-Vitorino F, Romaguera J, Zhao C, Vargas-Robles D, Ortiz-Morales G, Vázquez-Sánchez F, et al. Cervicovaginal Fungi and Bacteria Associated With Cervical Intraepithelial Neoplasia and High-Risk Human Papillomavirus Infections in a Hispanic Population. Front Microbiol. 2018;9:2533.

12. Castle PE, Hillier SL, Rabe LK, Hildesheim A, Herrero R, Bratti MC, et al. An association of cervical inflammation with high-grade cervical neoplasia in women infected with oncogenic human papillomavirus (HPV). Cancer Epidemiol Biomarkers Prev. 2001 Oct;10(10):1021–7.

13. Ravel J, Gajer P, Abdo Z, Schneider GM, Koenig SSK, McCulle SL, et al. Vaginal microbiome of reproductive-age women. Proc Natl Acad Sci U S A. 2011 Mar 15;108 Suppl 1:4680–7.

14. France MT, Ma B, Gajer P, Brown S, Humphrys MS, Holm JB, et al. VALENCIA: a nearest centroid classification method for vaginal microbial communities based on composition. Microbiome. 2020 Dec;8(1):166.

15. Godoy-Vitorino F, Romaguera J, Ortiz Morales G, Vázquez-Sánchez F, Dominicci-Maura A, Ortiz A. Malassezia globosa and Malassezia restricta are associated with high-risk HPV infections in the anogenital tract. Journal of Lower Genital Tract Disease. 2019 Apr 4;22:PMID: 30889031.

16. Chorna N, Romaguera J, Godoy-Vitorino F. Cervicovaginal Microbiome and Urine Metabolome Paired Analysis Reveals Niche Partitioning of the Microbiota in Patients with Human Papilloma Virus Infections. Metabolites. 2020 Jan 15;10(1):36.

17. Ilhan ZE, Łaniewski P, Thomas N, Roe DJ, Chase DM, Herbst-Kralovetz MM. Deciphering the complex interplay between microbiota, HPV, inflammation and cancer through cervicovaginal metabolic profiling. EBioMedicine. 2019 Jun;44:675–90.

18. Audirac-Chalifour A, Torres-Poveda K, Bahena-Román M, Téllez-Sosa J, Martínez-Barnetche J, Cortina-Ceballos B, et al. Cervical Microbiome and Cytokine Profile at Various Stages of Cervical Cancer: A Pilot Study. PLoS One. 2016;11(4):e0153274.

19. Güzel C, Govorukhina NI, Wisman GBA, Stingl C, Dekker LJM, Klip HG, et al. Proteomic alterations in early stage cervical cancer. Oncotarget. 2018 Apr 6;9(26):18128–47.

20. Olivier J, Johnson WD, Marshall GD. The logarithmic transformation and the geometric mean in reporting experimental IgE results: what are they and when and why to use them? Ann Allergy Asthma Immunol. 2008 Apr;100(4):333–7.

21. Moscicki AB, Shi B, Huang H, Barnard E, Li H. Cervical-Vaginal Microbiome and Associated Cytokine Profiles in a Prospective Study of HPV 16 Acquisition, Persistence, and Clearance. Frontiers in Cellular and Infection Microbiology [Internet]. 2020 [cited 2022 May 16];10. Available from: https://www.frontiersin.org/article/10.3389/fcimb.2020.569022

22. Scott ME, Shvetsov YB, Thompson PJ, Hernandez BY, Zhu X, Wilkens LR, et al. Cervical cytokines and clearance of incident human papillomavirus infection: Hawaii HPV cohort study: Mucosal cytokines and cervical HPV clearance. Int J Cancer. 2013 Sep 1;133(5):1187– 96.

23. Caporaso JG, Lauber CL, Walters WA, Berg-Lyons D, Huntley J, Fierer N, et al. Ultra-high-throughput microbial community analysis on the Illumina HiSeq and MiSeq platforms. ISME J. 2012 Aug;6(8):1621–4.

24. Gonzalez A, Navas-Molina JA, Kosciolek T, McDonald D, Vázquez-Baeza Y, Ackermann G, et al. Qiita: rapid, web-enabled microbiome meta-analysis. Nat Methods. 2018 Oct;15(10):796–8.

25. Amir A, McDonald D, Navas-Molina JA, Kopylova E, Morton JT, Zech Xu Z, et al. Deblur Rapidly Resolves Single-Nucleotide Community Sequence Patterns. Gilbert JA, editor. mSystems [Internet]. 2017 Apr 21 [cited 2022 Feb 24];2(2). Available from: https://journals.asm.org/doi/10.1128/mSystems.00191-16

26. Usyk M, Zolnik CP, Castle PE, Porras C, Herrero R, Gradissimo A, et al. Cervicovaginal microbiome and natural history of HPV in a longitudinal study. Silvestri G, editor. PLoS Pathog. 2020 Mar 26;16(3):e1008376.

27. DeSantis TZ, Hugenholtz P, Larsen N, Rojas M, Brodie EL, Keller K, et al. Greengenes, a Chimera-Checked 16S rRNA Gene Database and Workbench Compatible with ARB. Appl Environ Microbiol. 2006 Jul;72(7):5069–72.

28. Bolyen E, Rideout JR, Dillon MR, Bokulich NA, Abnet CC, Al-Ghalith GA, et al. Reproducible, interactive, scalable and extensible microbiome data science using QIIME 2. Nat Biotechnol. 2019 Aug;37(8):852–7.

29. Bray JR, Curtis JT. An Ordination of the Upland Forest Communities of Southern Wisconsin. Ecological Monographs. 1957 Oct;27(4):325–49.

30. Shannon CE. A Mathematical Theory of Communication. Bell System Technical Journal. 1948 Jul;27(3):379–423.

31. McMurdie PJ, Holmes S. phyloseq: An R Package for Reproducible Interactive Analysis and Graphics of Microbiome Census Data. PLOS ONE. 2013 Apr 22;8(4):e61217.

32. Hadley W. Ggplot2. New York, NY: Springer Science+Business Media, LLC; 2016.

33. Oksanen J, Simpson GL, Blanchet FG, Kindt R, Legendre P, Minchin PR, et al. vegan: Community Ecology Package [Internet]. 2022 [cited 2022 May 16]. Available from: https://CRAN.R-project.org/package=vegan

34. Anderson MJ. Permutation tests for univariate or multivariate analysis of variance and regression. Can J Fish Aquat Sci. 2001 Mar 1;58(3):626–39.

35. Segata N, Izard J, Waldron L, Gevers D, Miropolsky L, Garrett WS, et al. Metagenomic biomarker discovery and explanation. Genome Biol. 2011;12(6):R60.

36. Chong J, Liu P, Zhou G, Xia J. Using MicrobiomeAnalyst for comprehensive statistical, functional, and meta-analysis of microbiome data. Nat Protoc. 2020 Mar;15(3):799–821.

37. B Q, Z J, Q S, C J, Z L, X M. Cervicovaginal microbiota dysbiosis correlates with HPV persistent infection. Microbial pathogenesis [Internet]. 2021 Mar [cited 2022 Sep 15];152. Available from: https://pubmed.ncbi.nlm.nih.gov/33207260/

38. Saslow D, Solomon D, Lawson HW, Killackey M, Kulasingam SL, Cain J, et al. American Cancer Society, American Society for Colposcopy and Cervical Pathology, and American Society for Clinical Pathology screening guidelines for the prevention and early detection of cervical cancer. CA Cancer J Clin. 2012 Jun;62(3):147–72.

39. Vargas-Robles D, Magris M, Morales N, de Koning MNC, Rodríguez I, Nieves T, et al. High Rate of Infection by Only Oncogenic Human Papillomavirus in Amerindians. mSphere. 2018 Jun 27;3(3):e00176–18.

40. Castro FA, Quint W, Gonzalez P, Katki HA, Herrero R, van Doorn LJ, et al. Prevalence of and risk factors for anal human papillomavirus infection among young healthy women in Costa Rica. J Infect Dis. 2012 Oct 1;206(7):1103–10.

41. Geraets DT, Grünberg AW, van der Helm JJ, Schim van der Loeff MF, Quint KD, Sabajo LOA, et al. Cross-sectional study of genital carcinogenic HPV infections in Paramaribo, Suriname: prevalence and determinants in an ethnically diverse population of women in a pre-vaccination era. Sex Transm Infect. 2014 Dec;90(8):627–33.

42. Adebamowo SN, Ma B, Zella D, Famooto A, Ravel J, Adebamowo C. Mycoplasma hominis and Mycoplasma genitalium in the Vaginal Microbiota and Persistent High-Risk Human Papillomavirus Infection. Front Public Health. 2017 Jun 26;5:140.

43. Barut MU, Yildirim E, Kahraman M, Bozkurt M, Imirzalioğlu N, Kubar A, et al. Human Papilloma Viruses and Their Genotype Distribution in Women with High Socioeconomic Status in Central Anatolia, Turkey: A Pilot Study. Med Sci Monit. 2018 Jan 4;24:58–66.

44. Carozzi F, Puliti D, Ocello C, Anastasio PS, Moliterni EA, Perinetti E, et al. Monitoring vaccine and non-vaccine HPV type prevalence in the post-vaccination era in women living in the Basilicata region, Italy. BMC Infect Dis. 2018 Jan 15;18:38.

45. Grabowska AK, Riemer AB. The invisible enemy - how human papillomaviruses avoid recognition and clearance by the host immune system. Open Virol J. 2012;6:249–56.

46. Basith S, Manavalan B, Yoo TH, Kim SG, Choi S. Roles of toll-like receptors in cancer: a double-edged sword for defense and offense. Arch Pharm Res. 2012 Aug;35(8):1297–316.

47. Turner MD, Nedjai B, Hurst T, Pennington DJ. Cytokines and chemokines: At the crossroads of cell signalling and inflammatory disease. Biochim Biophys Acta. 2014 Nov;1843(11):2563–82.

48. Barros MR, de Oliveira THA, de Melo CML, Venuti A, de Freitas AC. Viral Modulation of TLRs and Cytokines and the Related Immunotherapies for HPV-Associated Cancers. J Immunol Res. 2018 May 2;2018:2912671.

49. Hu Z, Ma D. The precision prevention and therapy of HPV-related cervical cancer: new concepts and clinical implications. Cancer Med. 2018 Oct;7(10):5217–36.

50. Insinga RP, Perez G, Wheeler CM, Koutsky LA, Garland SM, Leodolter S, et al. Incident cervical HPV infections in young women: transition probabilities for CIN and infection clearance. Cancer Epidemiol Biomarkers Prev. 2011 Feb;20(2):287–96.

51. Sasagawa T, Takagi H, Makinoda S. Immune responses against human papillomavirus (HPV) infection and evasion of host defense in cervical cancer. J Infect Chemother. 2012 Dec;18(6):807–15.

52. Kim TK, Thomas SM, Ho M, Sharma S, Reich CI, Frank JA, et al. Heterogeneity of vaginal microbial communities within individuals. J Clin Microbiol. 2009 Apr;47(4):1181–9.

53. Bienkowska-Haba M, Luszczek W, Myers JE, Keiffer TR, DiGiuseppe S, Polk P, et al. A new cell culture model to genetically dissect the complete human papillomavirus life cycle. PLOS Pathogens. 2018 Mar 1;14(3):e1006846.

54. Tamarelle J, Thiébaut ACM, de Barbeyrac B, Bébéar C, Ravel J, Delarocque-Astagneau E. The vaginal microbiota and its association with human papillomavirus, Chlamydia trachomatis, Neisseria gonorrhoeae and Mycoplasma genitalium infections: a systematic review and meta-analysis. Clin Microbiol Infect. 2019 Jan;25(1):35–47.

55. Gillet E, Meys JFA, Verstraelen H, Verhelst R, De Sutter P, Temmerman M, et al. Association between Bacterial Vaginosis and Cervical Intraepithelial Neoplasia: Systematic Review and Meta-Analysis. PLoS One. 2012 Oct 2;7(10):e45201.

56. Brusselaers N, Shrestha S, van de Wijgert J, Verstraelen H. Vaginal dysbiosis and the risk of human papillomavirus and cervical cancer: systematic review and meta-analysis. Am J Obstet Gynecol. 2019 Jul;221(1):9–18.e8.

57. Lee JE, Lee S, Lee H, Song YM, Lee K, Han MJ, et al. Association of the vaginal microbiota with human papillomavirus infection in a Korean twin cohort. PLoS One. 2013;8(5):e63514.

58. Oh HY, Kim BS, Seo SS, Kong JS, Lee JK, Park SY, et al. The association of uterine cervical microbiota with an increased risk for cervical intraepithelial neoplasia in Korea. Clin Microbiol Infect. 2015 Jul;21(7):674.e1-9.

59. Reimers LL, Mehta SD, Massad LS, Burk RD, Xie X, Ravel J, et al. The Cervicovaginal Microbiota and Its Associations With Human Papillomavirus Detection in HIV-Infected and HIV-Uninfected Women. J Infect Dis. 2016 Nov 1;214(9):1361–9.

60. Fredricks DN, Fiedler TL, Marrazzo JM. Molecular Identification of Bacteria Associated with Bacterial Vaginosis. N Engl J Med. 2005 Nov 3;353(18):1899–911.

61. Cancer Facts & Figures for Hispanic/Latino People 2021-2023. :40.

62. Amir M, Brown JA, Rager SL, Sanidad KZ, Ananthanarayanan A, Zeng MY. Maternal Microbiome and Infections in Pregnancy. Microorganisms. 2020 Dec 15;8(12):E1996.

63. O’Hanlon DE, Moench TR, Cone RA. Vaginal pH and microbicidal lactic acid when lactobacilli dominate the microbiota. PLoS One. 2013;8(11):e80074.

64. Cruickshank R. The conversion of the glycogen of the vagina into lactic acid. The Journal of Pathology and Bacteriology. 1934;39(1):213–9.

65. Zhou Y, Wang L, Pei F, Ji M, Zhang F, Sun Y, et al. Patients With LR-HPV Infection Have a Distinct Vaginal Microbiota in Comparison With Healthy Controls. Front Cell Infect Microbiol. 2019;9:294.

66. Łaniewski P, Barnes D, Goulder A, Cui H, Roe DJ, Chase DM, et al. Linking cervicovaginal immune signatures, HPV and microbiota composition in cervical carcinogenesis in non-Hispanic and Hispanic women. Sci Rep. 2018 Dec;8(1):7593.

67. Campisciano G, Zanotta N, Licastro D, De Seta F, Comar M. In vivo microbiome and associated immune markers: New insights into the pathogenesis of vaginal dysbiosis. Sci Rep. 2018 Feb 2;8(1):2307.

68. De Seta F, Campisciano G, Zanotta N, Ricci G, Comar M. The Vaginal Community State Types Microbiome-Immune Network as Key Factor for Bacterial Vaginosis and Aerobic Vaginitis. Front Microbiol. 2019;10:2451.

69. Torcia MG. Interplay among Vaginal Microbiome, Immune Response and Sexually Transmitted Viral Infections. Int J Mol Sci. 2019 Jan 11;20(2):E266.

70. Filippi I, Carraro F, Naldini A. Interleukin-1β Affects MDAMB231 Breast Cancer Cell Migration under Hypoxia: Role of HIF-1α and NFκB Transcription Factors. Mediators Inflamm. 2015;2015:789414.

71. Apte RN, Voronov E. Is interleukin-1 a good or bad “guy” in tumor immunobiology and immunotherapy? Immunol Rev. 2008 Apr;222:222–41.

72. Iwata T, Fujii T, Morii K, Saito M, Sugiyama J, Nishio H, et al. Cytokine profile in cervical mucosa of Japanese patients with cervical intraepithelial neoplasia. Int J Clin Oncol. 2015 Feb;20(1):126–33.

73. Qian N, Chen X, Han S, Qiang F, Jin G, Zhou X, et al. Circulating IL-1beta levels, polymorphisms of IL-1B, and risk of cervical cancer in Chinese women. J Cancer Res Clin Oncol. 2010 May;136(5):709–16.

74. Arima K, Komohara Y, Bu L, Tsukamoto M, Itoyama R, Miyake K, et al. Downregulation of 15-hydroxyprostaglandin dehydrogenase by interleukin-1β from activated macrophages leads to poor prognosis in pancreatic cancer. Cancer Sci. 2018 Feb;109(2):462–70.

75. Guo Y, Xu F, Lu T, Duan Z, Zhang Z. Interleukin-6 signaling pathway in targeted therapy for cancer. Cancer Treat Rev. 2012 Nov;38(7):904–10.

76. Pahne-Zeppenfeld J, Schröer N, Walch-Rückheim B, Oldak M, Gorter A, Hegde S, et al. Cervical cancer cell-derived interleukin-6 impairs CCR7-dependent migration of MMP-9-expressing dendritic cells. International Journal of Cancer. 2014;134(9):2061–73.

77. Suradej B, Sookkhee S, Panyakaew J, Mungkornasawakul P, Wikan N, Smith DR, et al. Kaempferia parviflora Extract Inhibits STAT3 Activation and Interleukin-6 Production in HeLa Cervical Cancer Cells. International Journal of Molecular Sciences. 2019 Jan;20(17):4226.

78. Wei LH, Kuo ML, Chen CA, Chou CH, Lai KB, Lee CN, et al. Interleukin-6 promotes cervical tumor growth by VEGF-dependent angiogenesis via a STAT3 pathway. Oncogene. 2003 Mar;22(10):1517–27.

79. Wei LH, Kuo ML, Chen CA, Cheng WF, Cheng SP, Hsieh FJ, et al. Interleukin-6 in cervical cancer: the relationship with vascular endothelial growth factor. Gynecol Oncol. 2001 Jul;82(1):49–56.

80. Choi HS, Kim KM, Kim CH, Kim SM, Oh JS. Hydrogen Peroxide Producing Lactobacilli in Women with Cervical Neoplasia. Cancer Res Treat. 2006 Apr;38(2):108–11.

81. Vargas-Robles D, Morales N, Rodríguez I, Nieves T, Godoy-Vitorino F, Alcaraz LD, et al. Changes in the vaginal microbiota across a gradient of urbanization. Sci Rep. 2020 Dec;10(1):12487.

82. Eastment MC, McClelland RS. Vaginal microbiota and susceptibility to HIV. AIDS. 2018 Mar 27;32(6):687–98.

83. Chen Y, Qiu X, Wang W, Li D, Wu A, Hong Z, et al. Human papillomavirus infection and cervical intraepithelial neoplasia progression are associated with increased vaginal microbiome diversity in a Chinese cohort. BMC Infect Dis. 2020 Aug 26;20(1):629.

84. Anahtar MN, Byrne EH, Doherty KE, Bowman BA, Yamamoto HS, Soumillon M, et al. Cervicovaginal bacteria are a major modulator of host inflammatory responses in the female genital tract. Immunity. 2015 May 19;42(5):965–76.

85. Harwich MD, Serrano MG, Fettweis JM, Alves JMP, Reimers MA, Vaginal Microbiome Consortium (additional members), et al. Genomic sequence analysis and characterization of Sneathia amnii sp. nov. BMC Genomics. 2012;13 Suppl 8:S4.

86. Shipitsyna E, Roos A, Datcu R, Hallén A, Fredlund H, Jensen JS, et al. Composition of the vaginal microbiota in women of reproductive age--sensitive and specific molecular diagnosis of bacterial vaginosis is possible? PLoS One. 2013;8(4):e60670.

87. Caixeta RCA, Ribeiro AA, Segatti KD, Saddi VA, Fig.ueiredo Alves RR, dos Santos Carneiro MA, et al. Association between the human papillomavirus, bacterial vaginosis and cervicitis and the detection of abnormalities in cervical smears from teenage girls and young women. Diagn Cytopathol. 2015 Oct;43(10):780–5.

88. Boskey ER, Cone RA, Whaley KJ, Moench TR. Origins of vaginal acidity: high D/L lactate ratio is consistent with bacteria being the primary source. Hum Reprod. 2001 Sep;16(9):1809– 13.

89. Kroon SJ, Ravel J, Huston WM. Cervicovaginal microbiota, women’s health, and reproductive outcomes. Fertil Steril. 2018 Aug;110(3):327–36.

90. Libby EK, Pascal KE, Mordechai E, Adelson ME, Trama JP. Atopobium vaginae triggers an innate immune response in an in vitro model of bacterial vaginosis. Microbes Infect. 2008 Apr;10(4):439–46.

91. Vyshenska D, Lam KC, Shulzhenko N, Morgun A. Interplay between viruses and bacterial microbiota in cancer development. Semin Immunol. 2017 Aug;32:14–24.

92. Si J, You HJ, Yu J, Sung J, Ko G. Prevotella as a Hub for Vaginal Microbiota under the Influence of Host Genetics and Their Association with Obesity. Cell Host Microbe. 2017 Jan 11;21(1):97–105.

